# Establishing Fully-Automated Fundus-Controlled Dark Adaptometry: A Validation and Retest-Reliability Study

**DOI:** 10.1101/2023.06.09.23291212

**Authors:** Jeannine Oertli, Kristina Pfau, Hendrik P.N. Scholl, Brett G. Jeffrey, Maximilian Pfau

## Abstract

**Purpose:** To establish and validate a novel fundus-controlled dark-adaptometry method.

**Methods:** We developed a custom dark-adaptometry software for the S-MAIA device using the open perimetry interface. In the validation-substudy, participants underwent dark-adaptometry testing with a comparator device (MonCvONE, 59% rhodopsin bleach, cyan and red stimuli centered at 2°, 4°, and 6° eccentricity). Following a brief break (approx. 5 min), the participants were bleached again and underwent dark-adaptometry testing with the S-MAIA device (same loci). In the retest reliability-substudy, participants were tested twice with the S-MAIA device (same loci as above).

Nonlinear curve fitting was applied to extract dark-adaptation curve parameters. Validity and repeatability were summarized in terms of the mean bias and 95% limits of agreement (LoAs).

**Results:** In the validation-substudy (N=20 participants, median age [IQR] 31.5 years [25.8, 62.0]), measures of rod-mediated dark-adaptation showed little to no between method differences for the cone-rod-break-time (bias [95% CI] of +0.09 min [-0.5, 0.67]), rod-intercept-time (+0.42 min [-0.51, 1.35]), and S2 slope (-0.03 LogUnits/min [-0.04, -0.02]).

In the retest reliability-substudy (N=10 participants, 32.0 years [27.0, 57.5]), the corresponding LoAs were (cone-rod-break-time) -3.01 to 2.02 min, (rod-intercept-time) -3.95 to 2.94min, and (S2 slope) -0.09 to 0.07 LogUnits/min. The LoAs for the steady-state cone and rod thresholds were -0.27 to 0.31 LogUnits and -0.32 to 0.27 LogUnits.

**Conclusions:** The devised fundus-controlled dark-adaptometry method yields valid and reliable results. Fundus-controlled dark-adaptometry solves the critical need for localized testing of the visual cycle in eyes with unstable fixation (e.g., in the setting of subretinal gene therapy).

## INTRODUCTION

Early slowing of rod photoreceptor-mediated dark adaptation is characteristic for inherited retinal diseases (IRDs) due to Bruch’s membrane alterations,^1–5^ or enzymatic visual cycle dysfunction,^6, 7^ and for age-related macular degeneration (AMD),^8–11^ which constitutes the most common cause of legal blindness in industrialized countries.^12^ In these diseases, the ability to dark adapt following bright light exposure is typically impaired before fully dark-adapted rod sensitivity is lost (i.e., dynamic dysfunction preceding steady-state dysfunction).^3, 10, 13, 14^

Today, a plethora of methods are available for evaluating steady-state cone and rod function. Steady-state cone and rod dysfunction can be assessed using full-field electroretinography (retina-wide sum), and in a spatially-resolved manner using light- and dark-adapted two-color perimetry,^15, 16^ or chromatic pupil campimetry.^17^ Recently, fundus-controlled dark-adapted two-color perimetry (’microperimetry’) became available, enabling spatially-resolved testing of rod function even in patients with unstable fixation.^18–23^ In contrast, the widespread AdaptDx adaptometer (MacuLogix, Harrisburg, USA) is not fundus-controlled and only allows for large stimulus testing centered on eccentric loci (5° or 12°).^24, 25^ Free-viewing perimeters have two major drawbacks for evaluating patients following subretinally administered gene therapies: They are not suitable for assessing patients with unstable fixation, nor for assessing specific regions of interest in a patient-tailored manner.

Wadim Bowl and coworkers have previously used the MP1 microperimetry device for fundus-controlled dark adaptometry.^26, 27^ However, due to the limited dynamic range of the device, this method necessitated adding optical filters and changing the stimulus size within test runs (to accommodate for the limited dynamic range). Additionally, the workflow was also not fully automated, hindering its application in multicenter therapeutic trials.

To address this unmet need, we have now devised a fundus-controlled dark-adaptometry method for multicenter trials using a scanning laser ophthalmoscopy (SLO)-based microperimetry device (S-MAIA, CenterVue, Padova, Italy). As a prerequisite to the clinical application, we have evaluated the concurrent validity (against a commercially-available device), test-retest reliability, and construct validity of the devised method.

## METHODS

### Participants

This study was approved by the ethics committee for Northwestern and Central Switzerland (EKNZ) and adhered to the Declaration of Helsinki. Participants were informed of the study procedures and provided written informed consent before participating.

To be included, participants had to be older than 18 years and have no history of prior ocular surgery that - according to the investigator’s judgment - may affect visual function assessments (exceptions: cataract surgery, YAG laser capsulotomy, or laser retinopexy). Participants were initially enrolled in the validation substudy and subsequently in the test-retest reliability substudy. Participants could be enrolled in both substudies.

### Core Exams

All participants underwent autorefraction followed by best-corrected visual acuity testing (BCVA) using the qVA protocol (Manifold platform, Adaptive Sensory Technology, Lübeck, Germany). After the psychophysical tests (described below), the participants underwent SD-OCT imaging (30°x25°, 121 B-scans, ART 25, and BMO scan, Heidelberg Spectralis OCT2, Heidelberg Engineering, Heidelberg, Germany) to exclude any retinal or optic nerve head disease.

### Between-Device Validation Substudy

Following pupil dilatation, participants in the validation substudy (enrollment target N=20) underwent dark adaptometry testing with the MonCvONE device (Metrovision, Perenchies, France). The participants were bleached with the pre-set full-field 634 photopic cd/m^2^ (946 scotopic cd/m^2^) bleach for 5 minutes, corresponding to a 59% rhodopsin bleach.^13, 28^ Following the bleach, cyan and red Goldmann V-sized stimuli (peak wavelengths of 500 nm and 647 nm, stimulus duration 200 ms) were presented with 2°, 4°, and 6° eccentricity to the fovea. We selected these locations to explore whether rod-mediated dark adaptation can be examined in a clinical setting next to the fovea (given that psychophysical data,^13, 25^ and histopathologic data,^29^ suggest that AMD-associated rod dysfunction and degeneration is more severe toward the boundary of the rod-free zone).

We tested either along the nasal or temporal (horizontal) meridian (selected randomly). The thresholds were continuously determined for 30 minutes using the pre-set ’5-up-1-down staircase’ strategy. The examiner was allowed to terminate the test earlier if the final rod threshold was reached before 30 minutes.

For the newly devised method, we developed a custom *R* package utilizing the *open perimetry interface (OPI)* to communicate with the S-MAIA device,^30, 31^ and *Shiny* to provide a graphical user interface through a web application.^32^ The dark adaptation testing software is available as an *R* package (https://github.com/maximilianpfau *[Link-TBD]* [GNU GPLv3 license]).

Using the OPI workflow for the S-MAIA device, we first acquired the baseline fundus image and performed the brief initial fixation exam (approx. 10 seconds). Then, the participant was bleached with the MonCvONE device using the bleach specified above. After the 5-minute bleach, the participant had to position himself or herself in front of the S-MAIA device again quickly, and the fundus-controlled stimulus presentation started. Again, cyan and red stimuli (peak wavelengths: 505 nm and 627 nm; size: Goldmann III, duration: 200 ms) were presented with 2°, 4°, and 6° eccentricity to the fovea. The laterality (nasal/temporal retina) was matched with the MonCvONE test. A 5-up-1-down staircase strategy was used to obtain each threshold. The tests ran for 30 minutes. The examiner was allowed to terminate the test earlier if the final rod thresholds were reached.

### Test-Retest Reliability Substudy

In another 10 participants, we performed dark adaptation measurements with the devised S-MAIA-based method twice to determine the test-retest reliability of the devised method. Again, the stimuli could be presented nasally or temporally along the horizontal meridian, but the position within each participant was identical for both tests.

### Curve Fitting

For *<u>dark</u>* adaptation curve *<u>est</u>*imation (’darkest’), we developed a wrapper package in *R* using the *R* package *brms* for nonlinear curve fitting.^33^ The software is available at: https://github.com/maximilianpfau/ [Link-TBD] (GNU GPLv3 license).

For the dark adaptation curves from the MonCvONE, we used a regression formula describing both the cone and rod-mediated dark adaptation (modified from Flynn et al.).^13^

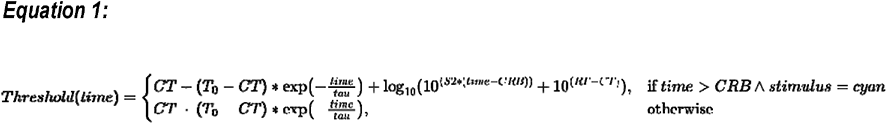

where

Threshold(time) = threshold (LogUnits) at a time (min)

T_0_ = initial threshold (LogUnits)

tau = exponential cone recovery time constant (min^-1^)

CT = cone threshold (LogUnits)

CRB = cone-rod break time (min)

S2 = S2 slope (LogUnits/min)

RT = final rod threshold (LogUnits)

We applied weakly informative priors for curve fitting that spanned the plausible value range for the curve parameters (Supplementary Table S1).

For the dark adaptation curves from the S-MAIA device, we used a regression formula representing only the rod-mediated dark adaptation phase since the first phase was not recorded (due to the time required to move the participant to the S-MAIA device after the bleach).

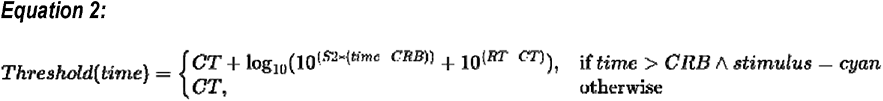

Data from the red stimuli were included in the fitting procedure up to <12 minutes. We excluded later red stimulus data from the curve fitting since eventual rod-meditation of the red stimulus was evident in most participants (cf., Figure 1).

**Figure 1.**
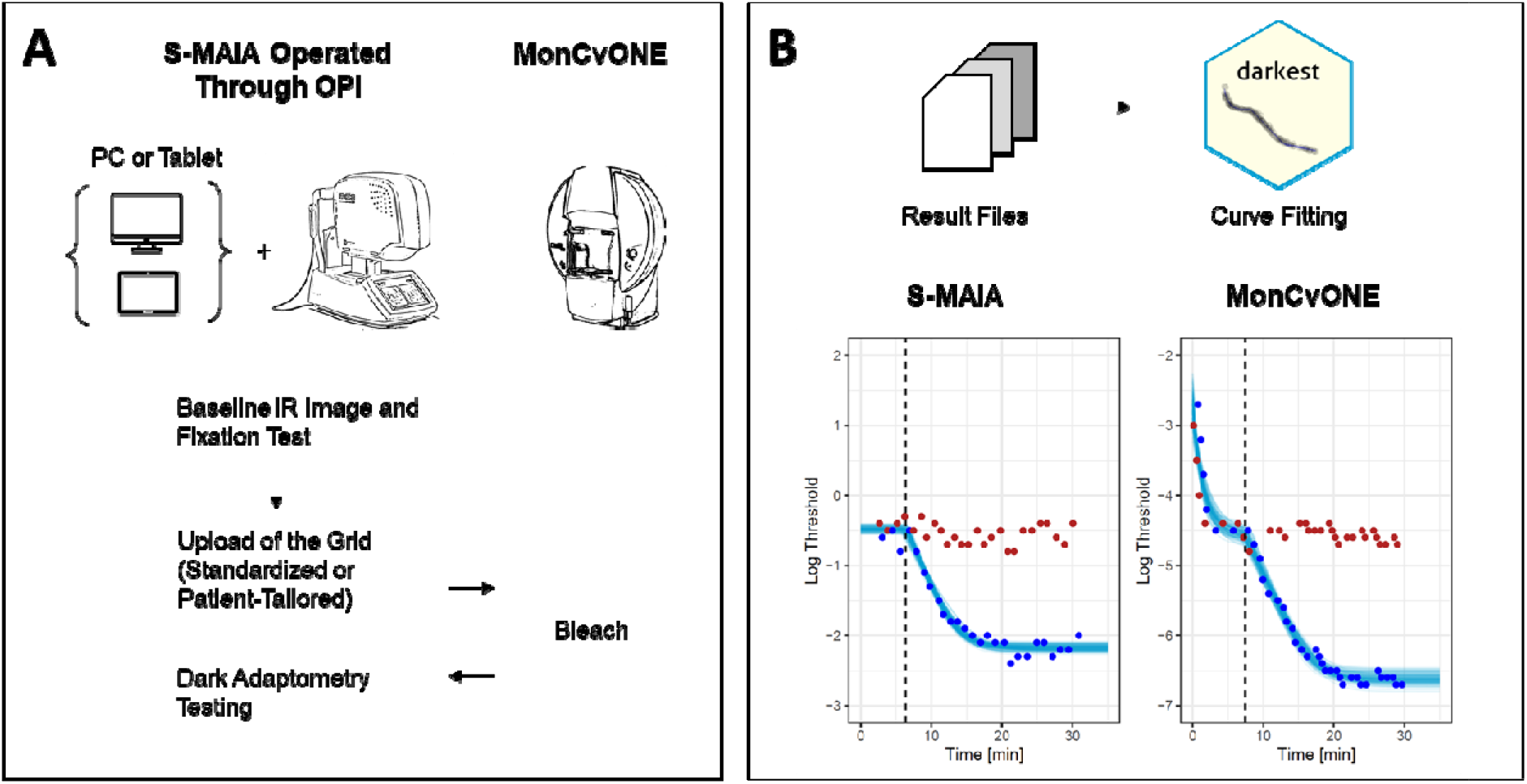
Dark-Adaptation with the S-MAIA Device. **Panel A** shows the workflow for the devised S-MAIA-based fundus-controlled dark adaptometry testing. In brief, participants were positioned in front of the S-MAIA device for a baseline infrared image and a short fixation test. Subsequently, the grid was uploaded to the custom software. Participants were then exposed to a background light that bleached the retina. Once the bleach was finished, the participants quickly returned to the S-MAIA device for threshold testing. **Panel B** shows typical dark adaptation curves (at 6°). Of note, the cone adaptation was typically not resolved with the S-MAIA-based testing due to the time required to switch from the MonCvONE device to the S-MAIA device. The red and blue dots denote the threshold for red and cyan stimuli. The semi-transparent lines show draws from the expectation of the posterior predictive distribution from the Bayesian nonlinear curve fit. The dashed vertical line denotes the cone-rod break time.

In addition, we extracted the rod intercept time (RIT) defined as the time to reach a criterion threshold of 1 LogUnit below the cone threshold (i.e., based on our average 6° cone threshold estimates across participants, the criterion thresholds were defined as -5.9 LogUnits for the MonCvOne and -1.4 LogUnits for the S-MAIA device).

To facilitate the comparison of the threshold estimates, we subtracted 4.3 LogUnits from MonCvONE-based measurements accounting for the difference in the dB-scales between the devices.

### Statistical Analysis

All statistical analyses were performed in the software environment *R* (data available at https://zenodo.org/ *[Link-TBD]*; analysis code available at: https://github.com/maximilianpfau *[Link-TBD]*).

BCVA and age were summarized based on their median and interquartile range. Data from participants tested at the temporal or nasal retina were pooled, given the absence of relevant differences (in relation to the retest reliability).

First, we used Bland-Altman plots for the (1.) between-method comparison and (2.) retest reliability analysis. For the analysis stratified by eccentricity, we used an intercept-only model to describe the measurement difference in terms of the mean bias (with the 95% confidence interval) and the 95% prediction intervals as the limits of agreement. For the joint analysis of the data from the three eccentricities, we fitted an intercept-only mixed model with the participant as a random effect (accounting for the repeated measures within each participant).

To analyze the association between age and the dark adaptation curve parameters, we fitted mixed models with the given parameter as the dependent variable, age and eccentricity as independent variables, and participant as a random effect.

## RESULTS

### Between-Method Comparison Substudy

A total of 23 participants were enrolled in the validation cohort. Of those, 20 were included in the between-method validation analysis (median age [IQR] 31.5 years [25.8, 62.0], Table 1).

**Table 1.**
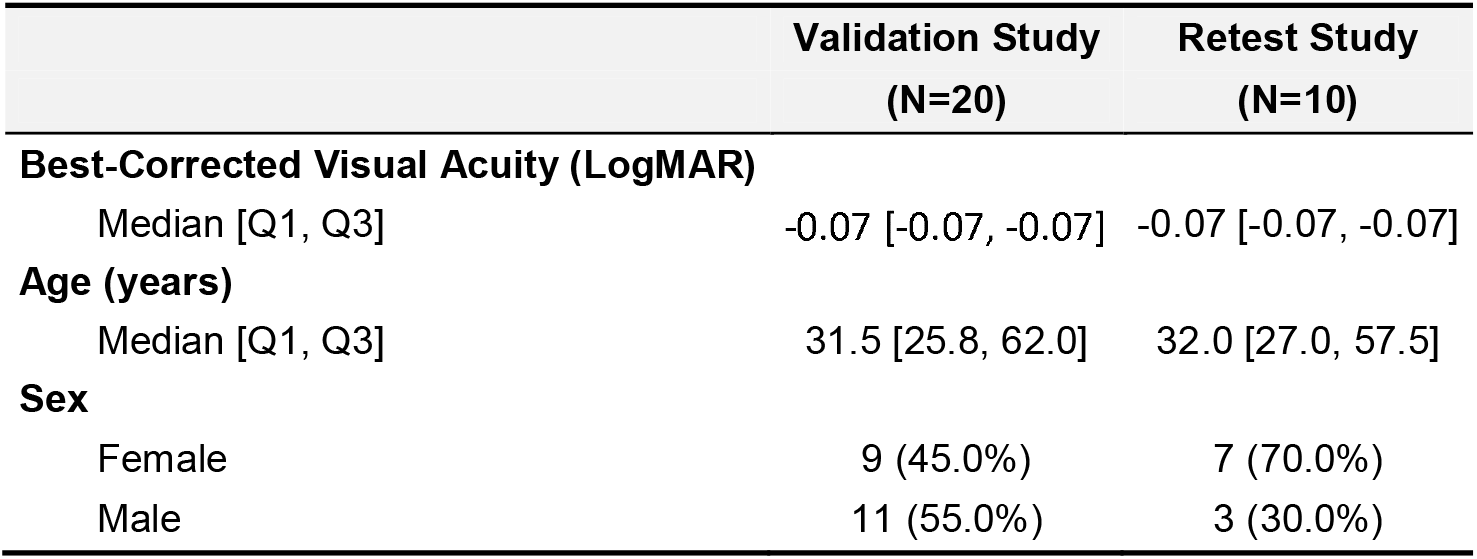
Participant Characteristics

|  | Validation Study<br>(N=20) | Retest Study<br>(N=10) |
| --- | --- | --- |
| **Best-Corrected Visual Acuity (LogMAR)** |  |  |
| Median [Q1, Q3] | -0.07 [-0.07, -0.07] | -0.07 [-0.07, -0.07] |
| **Age (years)** |  |  |
| Median [Q1, Q3] | 31.5 [25.8, 62.0] | 32.0 [27.0, 57.5] |
| **Sex** |  |  |
| Female | 9 (45.0%) | 7 (70.0%) |
| Male | 11 (55.0%) | 3 (30.0%) |

One participant was excluded from the analysis due to intermediate AMD, evidenced by multimodal imaging at the end of the study visit. Two participants were excluded from the analysis, since the laterality (nasal/temporal retina) was inadvertently flipped between the devices.

The MonCvONE and S-MAIA-based measurements yielded similar estimates for the (dynamic) measure of dark adaptation (Table 2, Figure 2). Specifically, there was no significant bias (MonCvONE -S-MAIA [95% CI]: +0.09 min [-0.5, 0.67]) for the cone-rod break with LoAs of -3.02 min and 3.19 min. Likewise, there was no significant bias for the rod intercept time (bias [95% CI] of +0.42 min [-0.51, 1.35]; LoAs of -4.39 min and 5.22 min).

**Figure 2.**
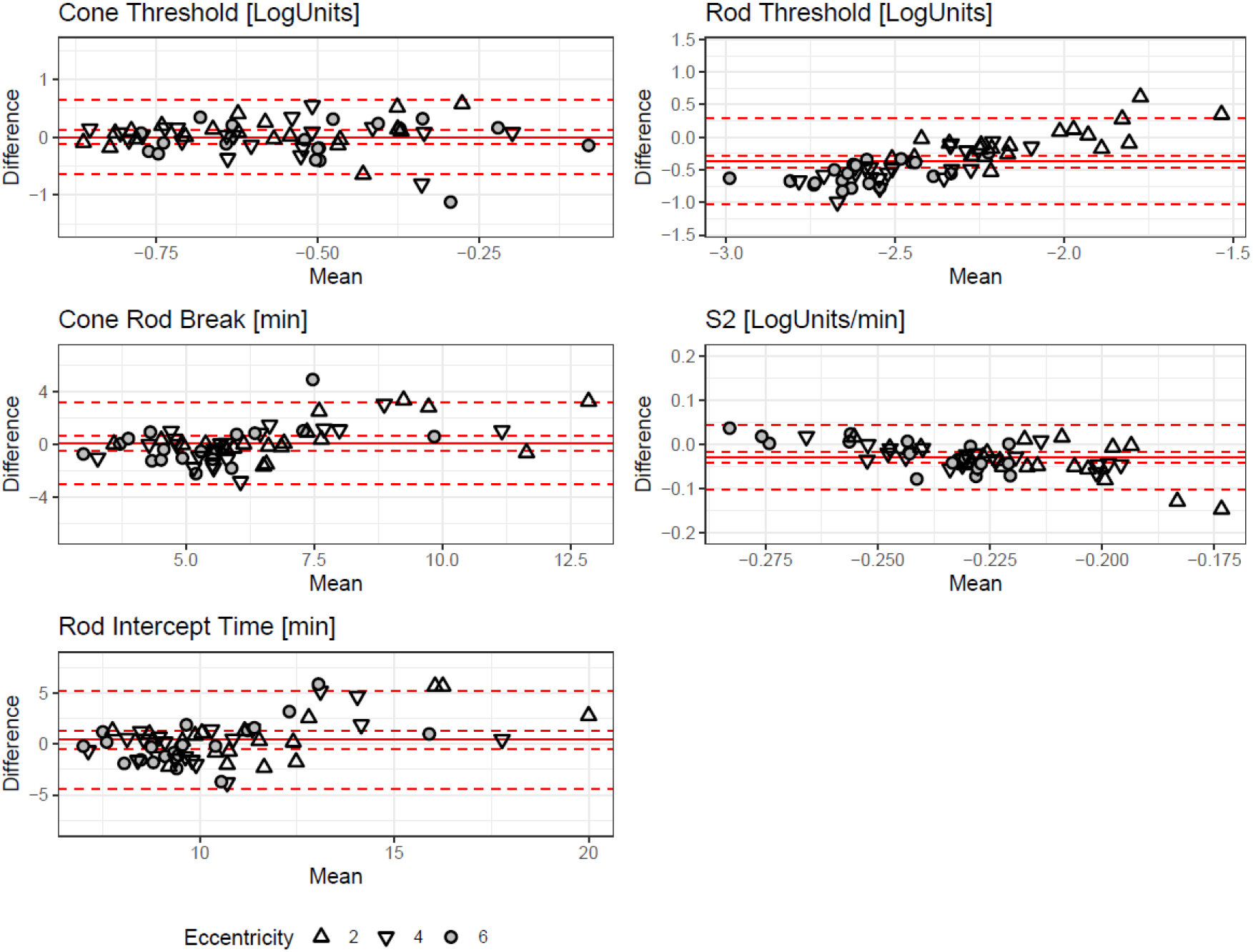
Between-Method Comparison. The Bland-Altman (mean-difference) plots show the between-method differences (MonCvONE minus S-MAIA) for the five parameters from the dark adaptation curve fitting. The shape denotes eccentricity (see legend). The solid red lines show the mean bias, the dashed inner lines the 95% confidence interval, and the dashed outer lines the 95% prediction intervals (limits of agreement).

**Table 2.** Between-Device Differences in Dark Adaptation Curve Parameters

| Parameter | Unit | Bias* [95% CI] | Lower LoA | Upper LoA |
| --- | --- | --- | --- | --- |
| Cone Rod Break Time (CRB) | Min | 0.09 [-0.5, 0.67] | -3.02 | 3.19 |
| Rod Intercept Time (RIT) | Min | 0.42 [-0.51, 1.35] | -4.39 | 5.22 |
| S2 Slope | LogUnits per Min | -0.03 [-0.04, -0.02] | -0.1 | 0.04 |
| Cone Threshold | LogUnits | 0 [-0.12, 0.12] | -0.65 | 0.65 |
| Rod Threshold | LogUnits | -0.37 [-0.46, -0.28] | -1.03 | 0.29 |

\* The values represent the MonCvONE minus S-MAIA difference.
*Abbreviations: Limit of Agreement (LoA)*

In contrast, the S2 slope was slightly steeper (i.e., more negative) in MonCvONE-based dark adaptation curves (bias [95% CI] of -0.03 LogUnits/min [-0.04, -0.02], LoAs of -0.1 LogUnits/min to 0.04 LogUnits/min).

The steady-state (threshold) parameters showed (*after accounting for the instrument-specific dB scales*) excellent agreement for the cone threshold with a bias of 0 LogUnits [-0.12, 0.12] and LoAs of -0.65 LogUnits and 0.65 LogUnits. The final rod thresholds were significantly lower for the MonCvONE-based measurements with a bias of -0.37 LogUnits [-0.46, -0.28] and LoAs of -1.03 LogUnits and 0.29 LogUnits.

The estimated curve parameters are shown in Supplementary Table S2. The between-method differences were overall similar across eccentricities (Supplementary Table S3).

### Test-Retest Reliability Substudy

Ten participants were enrolled in the intra-session test-retest reliability cohort (median age [IQR] of 32.0 years [27.0, 57.5], Table 1).

Overall, the intra-session test-retest reliability estimates showed no significant biases (i.e., no evidence of learning or fatigue effects), and the reliability was similar across all value ranges (Table 3, Figure 3).

**Figure 3.**
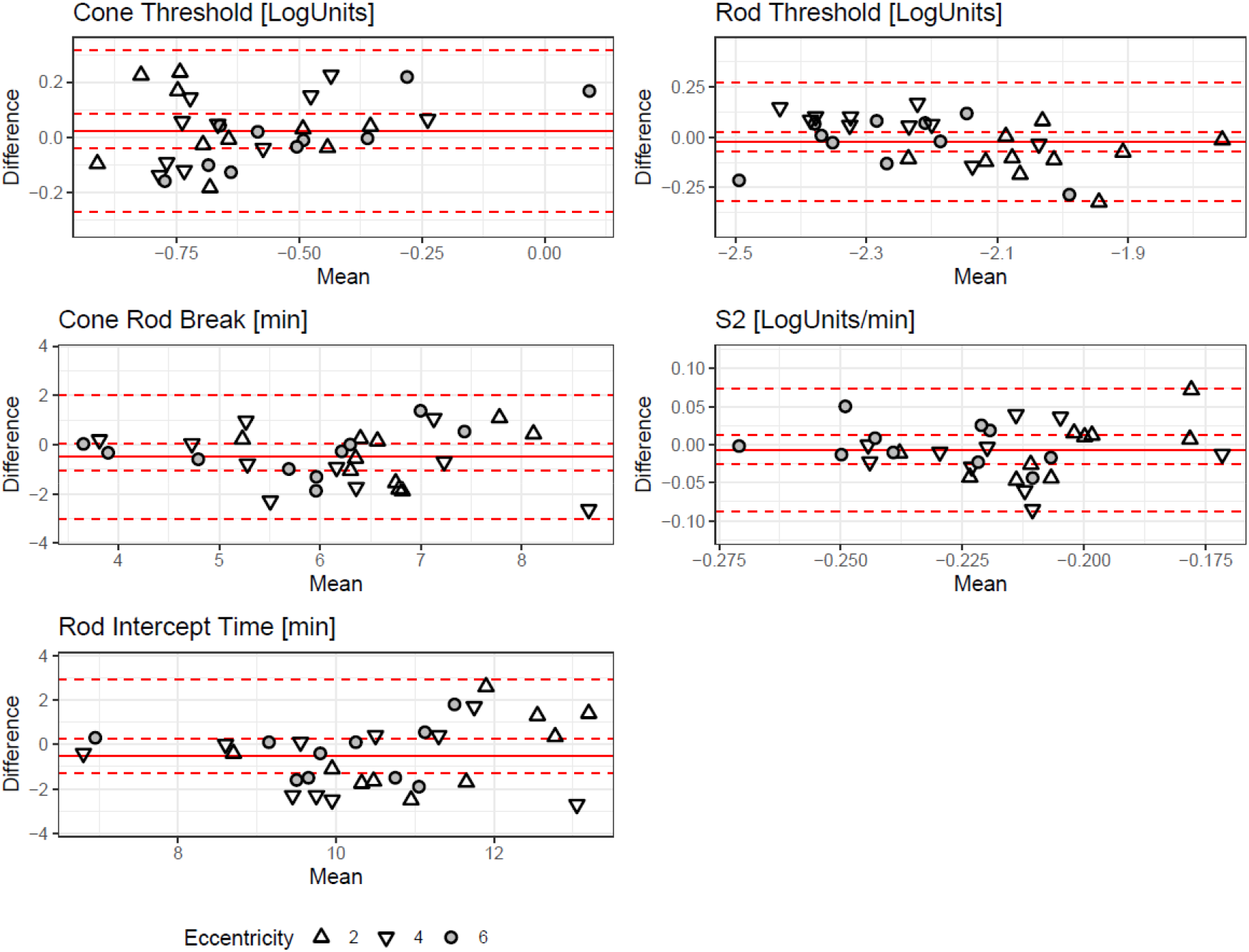
Test-Retest Reliability. The Bland-Altman (mean-difference) plots show the test-retest reliability for the five parameters from the dark adaptation curve fitting. The shape denotes eccentricity. The solid red lines show the mean bias, the dashed inner lines the 95% confidence interval, and the dashed outer lines the 95% prediction intervals (limits of agreement).

**Table 3.** Test-Retest Reliability of the Dark Adaptation Curve Parameters

| Parameter | Unit | Bias* [95% CI] | Lower LoA | Upper LoA |
| --- | --- | --- | --- | --- |
| Cone Rod Break Time (CRB) | Min | -0.49 [-1.05, 0.06] | -3.01 | 2.02 |
| Rod Intercept Time (RIT) | Min | -0.5 [-1.27, 0.26] | -3.95 | 2.94 |
| S2 Slope | LogUnits per Min | -0.01 [-0.03, 0.01] | -0.09 | 0.07 |
| Cone Threshold | LogUnits | 0.02 [-0.04, 0.08] | -0.27 | 0.31 |
| Rod Threshold | LogUnits | -0.02 [-0.07, 0.03] | -0.32 | 0.27 |

\* The values represent the first minus second test difference.
*Abbreviations: Limit of Agreement (LoA)*

The bias [95% CI] for the cone-rod break was -0.49 min [-1.05, 0.06] with LoAs of -3.01 min and 2.02 min, and for the rod intercept time -0.5 min [-1.27, 0.26] with LoAs of -3.95 min and 2.94 min. The bias for the S2 slope was -0.01 LogUnits/min [-0.03, 0.01] with LoAs of -0.09 LogUnits/min and 0.07 LogUnits/min.

For the steady-state (threshold) parameters, the test-retest reliability was excellent. For the cone threshold, the bias was 0.02 LogUnits [-0.04, 0.08] with LoAs of -0.27 LogUnits and 0.31 LogUnits, for the final rod threshold -0.02 LogUnits [-0.07, 0.03] with LoAs of -0.32 LogUnits and 0.27 LogUnits.

The test-retest reliability was similar across eccentricities (Supplementary Table S4).

### Association with Age

All S-MAIA-based data (validity and retest cohorts) from a total of 26 participants were pooled to assess the association of age with the curve parameters. All dark adaptation parameters increased with age, except for the cone threshold and S2 slope (Figure 4, Supplementary Table S5).

**Figure 4.**
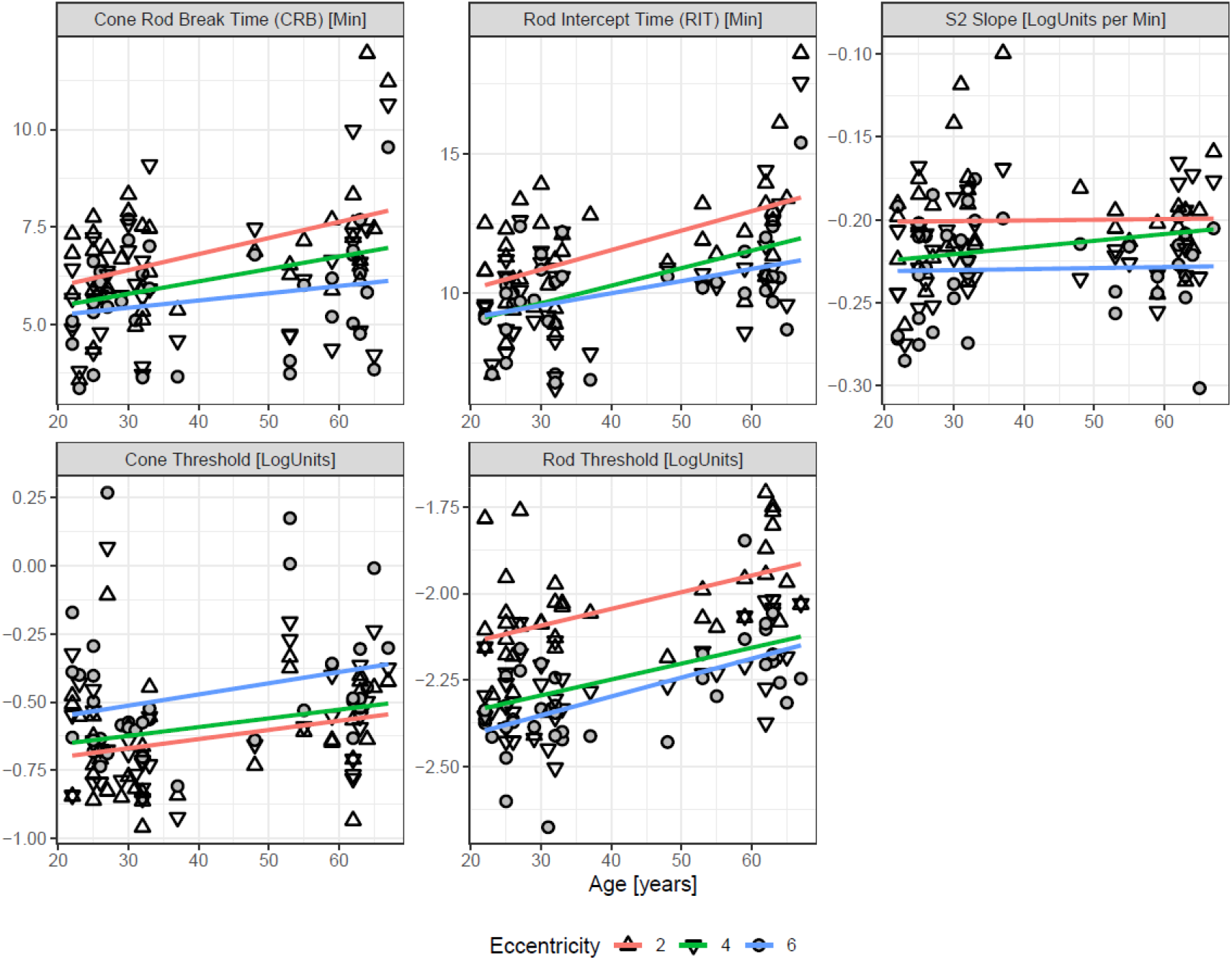
Association of Age with Dark Adaptation Parameters. The panels plot the dark adaptation curve parameters as a function of age. The shape denotes eccentricity. The regression lines are individual linear regression lines fitted for each eccentricity. Notably, the changes with age are similar across the three tested locations (i.e., no pronounced interaction effect between age and eccentricity).

The cone thresholds followed the hill-of-vision with slightly lower thresholds (i.e., better sensitivity) at 2° (intercept estimate of -0.77 LogUnits) compared to 4° and 6° (+0.04 LogUnits and +0.16 LogUnits). The change in cone sensitivity was not significant (slope estimate [95% CI] 0.04 LogUnits per decade [-0.01, 0.09], P=0.139).

In contrast, the rod threshold decreased with eccentricity (intercept of -2.22 LogUnits; effect estimates of -0.2 LogUnits for 4°, and -0.25 LogUnits for 6° eccentricity). The rod thresholds increased by (slope estimate [95% CI]) 0.04 LogUnits per decade [0.02, 0.06] (P<0.001).

The rod intercept time – as a measure of dynamic function – increased by 0.61 min per decade [0.22, 1.01] (P=0.002).

## DISCUSSION

Slowing of rod-mediated dark adaptation is among the earliest visual function alterations in AMD,^8–11^ and in IRDs due to Bruch’s membrane alterations,^1–5^ or enzymatic visual cycle dysfunction.^6, 7^ However, previous and ongoing clinical studies focused predominantly on large stimulus testing (Goldmann V or larger).^24, 25^ The now-devised method offers a fully-automated workflow for fundus-controlled dark adaptometry testing for clinical studies in patients with unstable fixation.

Critical advantages of the devised workflow are: (1) fundus tracking to examine patients with unstable fixation, (2) two-color testing to distinguish rod from cone mediation, (3) providing a fully-automated software for testing and analysis, (4) offering flexibility and precision for stimulus placement. Especially in the era of localized treatment (e.g., subretinally delivered gene therapies), this workflow provides the opportunity to test changes in function over time at specific regions of interest in a highly-localized manner. Analogously to the previous concept of patient-tailored perimetry,^34–36^ the devised workflow will enable researchers to use adaptometry to track leading disease fronts instead of coarse ’retina-wide’ testing.

Previously, Wadim Bowl and coworkers proposed the only other protocol for fundus-controlled dark adaptometry.^26, 27^ However, due to limitations of the MP1 microperimeter, their protocol necessitated adding filters to the optical path and changing the stimulus size during the exam to compensate for the low dynamic range. In addition, each test run had to be initiated manually, and no inter-device validation or retest reliability data was published for their method.^26^ To overcome these limitations, we developed a new method based on the S-MAIA device.

We selected as hardware the widely-available S-MAIA microperimetry device, which is widely distributed due to large multicenter studies such as MACUSTAR.^37^ Since the device can be operated through OPI,^30, 31^ a graphical user interface through a web application could be developed using *R Shiny*.^32^ The dark adaptation application is now available for non-technical users and provides flexibility regarding the testing grid and stimulus color (i.e., cyan and red testing versus only cyan testing).

As a critical prerequisite for the clinical application, this study established the concurrent validity, retest reliability, and construct validity of the devised method. The (dynamic) dark adaptation metrics (cone-rod break, RIT, and S2 slope) were concordant among the comparator and the devised method. Importantly, the minor between-device biases of these metrics were markedly smaller than the retest variability,^38^ and typical disease-associated changes.^10^

The steady-state parameters (cone threshold) were – accounting for the instrument-specific difference in the dB-scale – in agreement as indicated by the tight limits of agreement. Interestingly, the absolute differences for the cone minus rod thresholds were larger in MonCvONE-than in S-MAIA-based measurements (e.g., at 6° 2.3 vs. 1.8 LogUnits, cf., Supplementary Table S2). This difference is most likely attributable to two stimulus size-related factors: First, the fundus-controlled Goldmann III stimuli (diameter of 0.43° or 125 µm) will stimulate to a lesser degree, rod photoreceptors distant to the locus center compared to a Goldmann V stimuli (diameter of 1.72° or 500 µm). This can affect rod thresholds given that all loci were within the region where rod photoreceptor density increases steeply with eccentricity.^39^ Second, the perception of the Goldmann III stimulus is most likely affected by a shifting of the spatial summation curves during dark adaptation. While the Goldmann V stimulus exceeds Ricco’s area in photopic and scotopic spatial summation curves (in the central macula), the Goldmann III stimulus exceeds Ricco’s area under photopic conditions, but not under scotopic conditions.^40–43^ Thus, the S-MAIA-based cone thresholds are measured with complete (or near complete) spatial summation, while rod thresholds are measured along the ascending arm of the intensity-response function.

The retest reliability estimates were similar to previously reported data for all dark adaptation curve parameters, underscoring the devised method’s reliability.^38^

Our cone and rod thresholds match the photoreceptor distribution (i.e., lowest cone threshold at 2°, lowest final rod threshold at 6°).^39^ The steeper age-associated decline of rod-related function is compatible with histopathologic studies documenting severe age-associated rod loss between 0.5 and 3 mm eccentricity.^29^ Also, our estimates for the cone-rod break and RIT at 4° and 6° are – when accounting for age differences (cf., effect of age estimate in Supplementary Table S5) – similar to previous data in healthy adults and early AMD patients that used 50% to 70% rhodopsin bleaches.^13, 44, 45^

### Limitations

Our study was restricted to healthy volunteers, so it is yet to be determined how effective our methods will be in patients with retinal diseases. We hypothesize that fundus-controlled dark adaptometry will provide even greater benefits over free-viewing dark adaptometry in patients with suboptimal or unstable fixation. In addition, the sample size for the retest reliability estimates is small.

In summary, we devised a novel method for fundus-controlled dark adaptometry. Key advantages of the devised method are the wide availability of the microperimetry device, the ability to perform dark adaptometry in patients with unstable fixation, and the flexibility of the developed software enabling custom tests. Fundus-controlled dark adaptometry will facilitate monitoring localized changes in rod function in clinical trials for IRDs and AMD.

## Data Availability

All data produced in the present study are available upon reasonable request to the authors

